# Prior infection- and/or vaccine-induced protection against Omicron BA.1, BA.2 and BA.4/BA.5-related hospitalisations in older adults: a test-negative case-control study in Quebec, Canada

**DOI:** 10.1101/2022.12.21.22283740

**Authors:** Sara Carazo, Danuta M. Skowronski, Marc Brisson, Chantal Sauvageau, Nicholas Brousseau, Judith Fafard, Rodica Gilca, Denis Talbot, Manale Ouakki, Yossi Febriani, Geneviève Deceuninck, Philippe De Wals, Gaston De Serres

**Author notes:** **Corresponding author:** Sara Carazo, Institut national de santé publique du Québec, 2400, Avenue d’Estimauville, Québec, Québec, G1E 7G9, Canada.

## Abstract

**Background:** Due to severe outcomes, elderly adults 60 years or older are prioritized for COVID-19 vaccination but accumulated SARS-CoV-2 infection and vaccination likely modifies their risk. We estimated vaccine effectiveness against omicron-associated hospitalisation among elderly adults, by number of doses, prior infection history and time since last immunological event.

**Methods:** We conducted a test-negative case-control study among symptomatic elderly adults tested for SARS-CoV-2 in Quebec, Canada during BA.1-, BA.2- and BA.4/5-dominant periods. Relative to unvaccinated, infection-naïve participants, we compared COVID-19 hospitalisation risk by mRNA vaccine dose and/or prior infection (pre-omicron or omicron) history.

**Findings:** During BA.1, BA.2 and BA.4/5 periods, two- vs. four-dose vaccine effectiveness alone against hospitalisation was: 78% (95%CI:75-80) vs. 96% (95%CI:93-98); 60% (95%CI:50-97) vs. 84% (95%CI:81-87); and 40% (95%CI:30-49) vs. 68% (95%CI:63-72), respectively, consistent with longer median time since second vs fourth dose. By respective period, effectiveness of pre-omicron vs. omicron infection alone against hospitalisation was: 93% (95%CI:80-97) vs. [not estimable]; 88% (95%CI:50-97) vs. 96% (95%CI:68-99); and 69% (95%CI:30-85) vs. 90% (95%CI:79-95). Regardless of doses (2-5) or prior infection type, hybrid protection was ≥90%, lasting at least 6-8 months during the BA.4/5 period. Prior omicron infection alone reduced BA.4/5 hospitalisation risk by >80% for at least 6-8 months.

**Interpretation:** Elderly adults with history of both prior SARS-CoV-2 infection and ≥2 vaccine doses appear well-protected for a prolonged period against omicron hospitalisation, including BA.4/5. Ensuring infection-naïve older adults remain up-to-date with vaccination may further reduce COVID-19 hospitalisations most efficiently.

**Funding:** Ministère de la Santé et des Services sociaux du Québec.

## INTRODUCTION

Since December 2021, omicron (B.1.1.529) has been the dominant SARS-CoV-2 variant globally.^1^ The initial omicron BA.1 sublineage was displaced by the more transmissible and immune evasive BA.2, the latter succeeded thereafter by BA.4 and BA.5 sublineages that have since predominated through November 2022.^2^ BA.4 and BA.5 (BA.4/5) demonstrate substantial escape from vaccine or infection-induced neutralizing antibodies but this immune evasion is less pronounced in individuals with hybrid immunity resulting from both prior infection and vaccination.^3^

Vaccine effectiveness (VE) against SARS-CoV-2 is reduced and of shorter duration for omicron compared to the delta variant,^4–6^ and for the BA.4/5 omicron sublineage compared to BA.1 or BA.2.^7–9^ Stronger and longer-lasting VE during BA.1 and BA.2 dominant periods was reported among individuals with history of prior infection,^10,11^ but similar data for hybrid immunity against BA.4/5 are lacking, including for severe outcomes or by number of vaccine doses, type of prior infection (pre-omicron or omicron) or duration since the last immunological event.

Owing to their greater risk of severe outcomes, elderly adults ≥60 years of age have been prioritized for COVID-19 vaccination since the start of the pandemic. Sero-prevalence data in Canada, the United States, United Kingdom and elsewhere show age-related decrease in cumulative SARS-CoV-2 infection rates across the pandemic^12–15^ Infections foremost accrued during successive omicron waves, with at least 70% of Canadian children and young adults and about half of elderly adults having been infected by autumn 2022.^12,13^ The accumulating percentage of the elderly population with prior history of both SARS-CoV-2 infection and repeat vaccination has likely modified their baseline risk. However, most VE analyses and booster dose policies have not taken into account this relevant and evolving immuno-epidemiological context.^16–18^

The objective of this test-negative case-control study was to estimate the protection against omicron-associated hospitalisation among elderly adults during BA.1, BA.2 and BA.4/5 periods by number of vaccine doses (one to five), and/or prior infection history (pre-omicron or omicron) and time since last immunological event (vaccination or infection).

## METHODS

### Study design

This population-based test-negative case-control study included symptomatic elderly adults aged 60 years or older living in the province of Quebec, Canada who were tested for SARS-CoV-2 by nucleic acid amplification testing (NAAT) during an emergency room visit or upon hospital admission between December 26, 2021 and November 5, 2022.

The study was conducted under the legal mandate of the National Director of Public Health of Quebec under the Public Health Act and was also approved by the Research Ethics Board of the Centre hospitalier universitaire de Québec-Université Laval.

### Data sources

Source data included four Quebec population databases that were linked through a unique identifying number including: (1) the provincial laboratory database, comprising all individual-level NAAT details and results, including testing indication, since pandemic start; (2) the provincial immunisation registry, including all Quebec residents and their COVID-19 vaccine doses and dates administered since the immunization campaign began; (3) the administrative hospitalisation database; and (4) the provincial chronic disease surveillance database.

### Sublineage dominant periods

Between December 26, 2021 and March 13, 2022, the Quebec laboratory-based surveillance system used single nucleotide polymorphism genotyping of viruses collected from designated sentinel sites for the attribution of omicron lineage. From March 14, 2022, whole genome sequencing was conducted on a randomly-selected sample of all positive NAAT specimens across the province. Based on this viral genetic surveillance, three analysis periods were defined by predominant omicron sublineage: BA.1 (74-99% of characterized viruses weekly) spanning December 26, 2021 to March 12, 2022; BA.2 (74-99% of characterized viruses weekly) spanning April 3 to June 11, 2022; and BA.4/5 spanning July 3 to November 5, 2022 when BA.4 comprised 15-21% and BA.5 and descendant variants (including BQ.1.1) comprised 61-81% of characterized viruses weekly (**Figure 1**). BQ.1.1 sublineage detection increased from 4% to 15% during the final four weeks of the study. Weeks during which more than one sublineage co-circulated at high levels were excluded.

**Figure 1.**
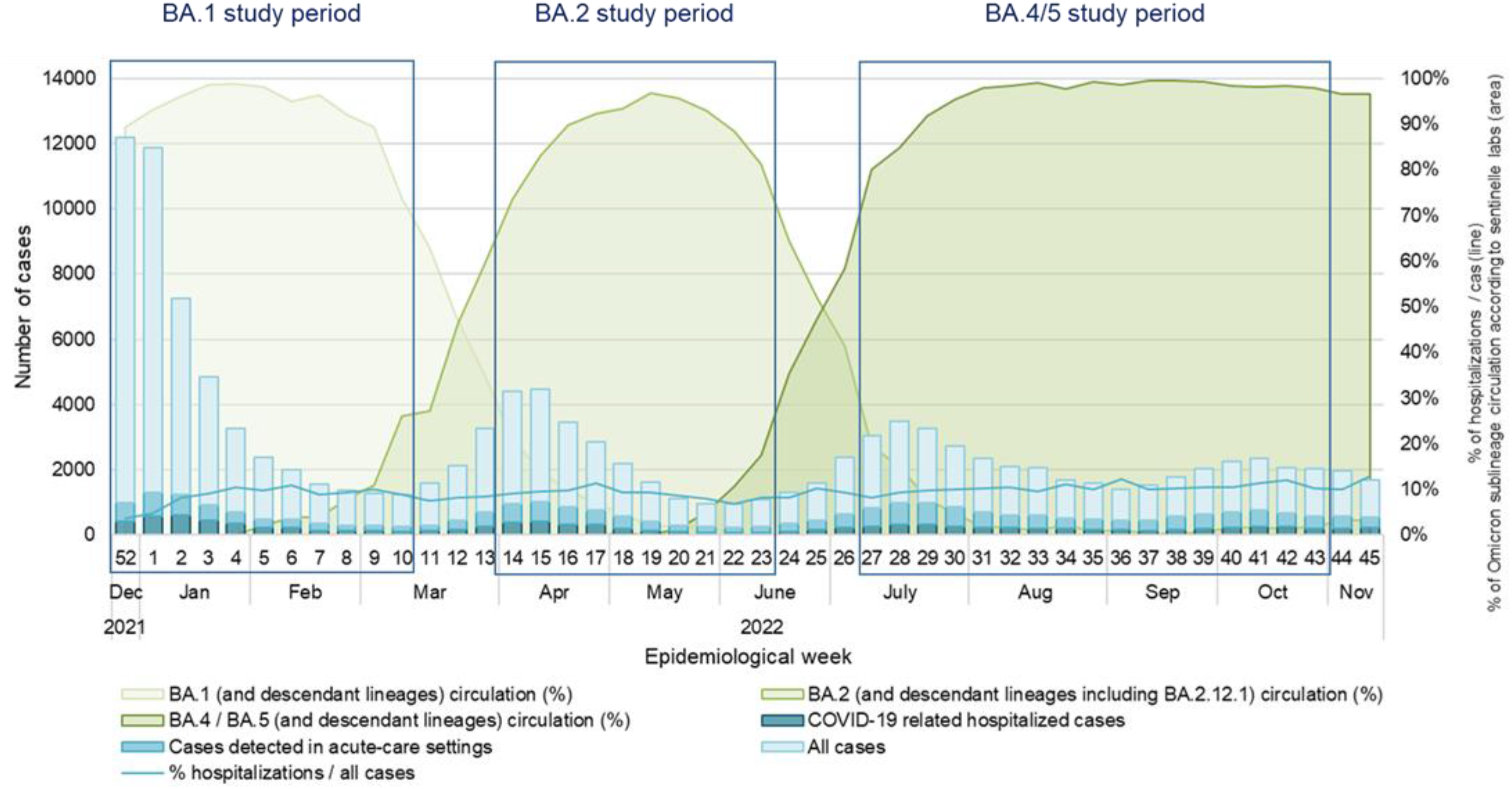
Number of weekly COVID-19 cases, emergency department consultations and hospitalisations among elderly adults 60 years or older and omicron sublineage circulation in the province of Quebec, Canada

We used similar dominance periods to ascribe the variant of prior infection, assigning the initially circulating variant/sublineage to weeks with cocirculation as follows: pre-omicron if detected before December 26, 2021; BA.1 from December 26, 2021 to April 2, 2022; BA.2 from April 3 to July 2, 2022 and BA.4/5 after July 3, 2022.

### Outcome and exposure definitions

In Quebec, NAAT access was restricted to prioritized groups and those with severe outcomes during the first pandemic wave but was made widely available thereafter from July 2020. With the substantial omicron surge contemporaneous with rapid antigen detection tests (RADT) being made broadly available free-of-charge to the general population, NAAT was restricted again from January 5, 2022 to individuals ≥70 years of age with COVID-19-compatible symptoms, patients consulting in emergency departments or admitted to hospital, and residents of closed settings.^19^

For the current study, eligible participants were symptomatic elderly adults tested for SARS-CoV-2 by NAAT at an emergency department. Hospitalised cases were those SARS-CoV-2 NAAT positive and admitted 0 to 14 days after specimen collection for respiratory illness or with COVID-19 as their main diagnosis at discharge. Controls were participants with a negative SARS-CoV-2 NAAT result, with or without subsequent hospitalisation.

Exposure was defined by a combination of prior infection and vaccination history. Vaccination was defined as the administration of 1-5 doses of BNT162b2 (Pfizer-BioNTech) or mRNA-1273 (Moderna) (mRNA) monovalent vaccines ≥14 days before specimen collection (for first dose) or ≥7 days before specimen collection (for second to fifth doses). An interval ≥21 days between first and second doses and ≥90 days between subsequent booster doses was required, as per provincial guidelines. Prior infection was defined by a first positive SARS-CoV-2 NAAT result ≥60 days before the current specimen collection. A 60-day interval was chosen to capture most reinfections, balancing improved sensitivity to detect early reinfections against imperfect specificity due to prolonged viral shedding, which may be more frequent among elderly adults.^20–22^ The less sensitive but more specific 90-day interval was also explored in sensitivity analyses.

### Exclusion criteria

We excluded specimens from individuals who lived in long-term care facilities (LTCF), had missing chronic disease data, had a documented reinfection (60-day interval) before the study period or had received >5 doses or a non-mRNA vaccine dose and those whose minimum intervals between doses or between vaccination and sampling were not respected. In sensitivity analyses we also excluded individuals identified as immunosuppressed.

### Statistical analyses

For each analysis period, logistic regression models estimated the odds ratio (OR) for omicron-associated hospitalisation by vaccine dose and/or prior infection history relative to unvaccinated, infection-naïve individuals. All negative tests were included as controls, but repeat tests conducted among cases were censored during the 59 days following their positive specimen.^23^ Models were adjusted for age (60-69, 70-79, 80-89 and ≥90 years old), sex, type of residence (home, private homes for elderly and other), presence of chronic respiratory disease, chronic heart disease, cancer, obesity, immunosuppressive condition, neurological disease, multimorbidity (defined as at least two conditions), and epidemiological week of sampling. Protection or VE was derived as (1 − OR) × 100. Incremental protection was also estimated comparing individuals who received 3-5 mRNA vaccine doses to those vaccinated with two doses ≥6 months earlier.

Protection was stratified by time since last immunological event: vaccine dose or primary infection. Previously infected individuals who received two to five doses were pooled to estimate protection by time since last vaccine dose given the few hospitalised cases and previous results showing similar protection by time since last dose, regardless of dose number.^9,11^

Statistical analyses were performed using SAS (version 9.4).

## RESULTS

There were 293,722 NAATs performed in total among elderly adults consulting emergency departments with COVID-19 compatible symptoms between December 26, 2021 and November 5, 2022, with 247,292 collected during specified omicron analysis periods. The proportion of excluded NAATs was similar during the BA.1, BA.2 and BA.4/5 periods (19,906 (26.5%), 15,010 (24.4%) and 24,094 (21.8%), respectively). The most frequent reason for exclusion was LTCF residence (n=15907 [6.4%]) followed by non-mRNA vaccination (n=15,116 [6.1%]) (**Supplementary figure 1**). After exclusions, there were 2951, 1897 and 3607 hospitalised cases and 48,724, 41,702 and 75,938 controls during the BA.1, BA.2 and BA.4/5 analysis periods, respectively.

In all periods, cases compared to controls were more often men (4637 [54.8%] vs 78,233 [47.0%]), aged ≥80 years (4405 [52.1%] vs 66,684 [40.1%]) and had at least two comorbid conditions (6919 [81.8%] vs 123,824 [74.4%]) (**Table 1**). Among controls, 7346 (4.4%) were unvaccinated, 77,812 (46.8%) received only Pfizer-BioNTech vaccine, 26,204 (15.8%) only Moderna, and 55,002 (33.1%) had a mixed Pfizer-BioNTech/Moderna schedule, a proportion that increased from 21.2% during the BA.1 period to 34.7% and 39.8% during the BA.2 and BA.4/5 periods, respectively.

**Table 1.** Characteristics of omicron hospitalised cases and test-negative controls stratified by omicron sublineage analysis period

|  | **BA.1 period**<br>n=51,675 |  | **BA.2 period**<br>n=43,599 |  | **BA.4/5 period**<br>n=86,600 |  |
| --- | --- | --- | --- | --- | --- | --- |
| **Characteristic** | **Cases**<br>(n=2951) | **Controls**<br>(n=48724) | **Cases**<br>(n=1897) | **Controls**<br>(n=41702) | **Cases**<br>(n=3607) | **Controls**<br>(n=75938) |
| **Sex** |  |  |  |  |  |  |
| Female | 1290 (43.7) | 25417 (52.2) | 869 (45.8) | 21989 (52.7) | 1659 (46.0) | 40725 (53.6) |
| Male | 1661 (56.3) | 23307 (47.8) | 1028 (54.2) | 19713 (47.3) | 1948 (54.0) | 35213 (46.4) |
| **Age, years** | 78 (71 – 86) | 77 (69 – 84) | 81 (75 – 88) | 76 (68 – 84) | 80 (73 – 87) | 77 (69 – 84) |
| 60-69 | 644 (21.8) | 13232 (27.2) | 254 (13.4) | 11812 (28.3) | 517 (14.3) | 20363 (26.8) |
| 70-79 | 931 (31.5) | 15612 (32.0) | 542 (28.6) | 13919 (33.4) | 1162 (32.2) | 24742 (32.6) |
| 80-89 | 968 (32.8) | 14568 (29.9) | 755 (39.8) | 11847 (28.4) | 1391 (38.6) | 22183 (29.2) |
| ≥90 | 408 (13.8) | 5312 (10.9) | 346 (18.2) | 4124 (9.9) | 537 (14.9) | 8650 (11.4) |
| **Origin** |  |  |  |  |  |  |
| Home | 2194 (74.3) | 36030 (73.9) | 1104 (58.2) | 32330 (77.5) | 2774 (76.9) | 57198 (75.3) |
| Private seniors' homes | 684 (23.2) | 11615 (23.8) | 755 (39.8) | 8527 (20.4) | 772 (21.4) | 17068 (22.5) |
| Other | 73 (2.5) | 1079 (2.2) | 38 (2.0) | 845 (2.0) | 61 (1.7) | 1672 (2.2) |
| **Comorbidity<sup>a</sup>** |  |  |  |  |  |  |
| At least two conditions | 2405 (81.5) | 37155 (76.3) | 1595 (84.1) | 30909 (74.1) | 2919 (80.9) | 55760 (73.4) |
| Chronic heart disease | 1578 (53.5) | 24279 (49.8) | 1077 (56.8) | 19513 (46.8) | 1896 (52.6) | 34938 (46.0) |
| Chronic lung disease | 1178 (39.9) | 17304 (35.5) | 798 (42.1) | 14831 (35.6) | 1379 (38.2) | 26560 (35.0) |
| Cancer | 640 (21.7) | 10713 (22.0) | 444 (23.4) | 8916 (21.4) | 792 (22.0) | 15277 (20.1) |
| Neurologic disease / dementia | 515 (17.5) | 6564 (13.5) | 386 (20.3) | 5598 (13.4) | 660 (18.3) | 10519 (13.9) |
| Obesity | 420 (14.2) | 5656 (11.6) | 228 (12.0) | 4799 (11.5) | 404 (11.2) | 8225 (10.8) |
| Immunosuppressive condition | 262 (8.9) | 3486 (7.2) | 148 (7.8) | 2910 (7.0) | 331 (9.2) | 5184 (6.8) |
| **Prior infection history** |  |  |  |  |  |  |
| No prior infection | 2918 (98.9) | 46323 (95.1) | 1845 (97.3) | 37292 (89.4) | 3440 (95.4) | 63927 (84.2) |
| Pre-omicron prior infection<br>(Mar 1, 2020 to Dec 25, 2021) | 32 (1.1) | 2322 (4.8) | 35 (1.8) | 1840 (4.4) | 51 (1.4) | 3012 (4.0) |
| Omicron prior infection<br>(Dec 26, 2021 to Aug 8, 2022) | 1 (0.0) | 79 (0.2) | 17 (0.9) | 2570 (6.2) | 116 (3.2) | 8999 (11.9) |
| Omicron BA.1 (Dec 26, 2021 to Apr 9, 2022) | 1 (0.0) | 79 (0.2) | 16 (0.8) | 2549 (6.1) | 69 (1.9) | 5316 (7.0) |
| Omicron BA.2 (Apr 10 to July 9, 2022) | NA | NA | 1 (0.1) | 21 (0.1) | 32 (0.9) | 2825 (3.7) |
| Omicron BA.4/5 (July 10 to Sep 6, 2022)<sup>b</sup> | NA | NA | NA | NA | 15 (0.4) | 858 (1.1) |
| **Vaccination status and type of vaccine** |  |  |  |  |  |  |
| Unvaccinated | 776 (26.3) | 2289 (4.7) | 221 (11.6) | 1847 (4.4) | 330 (9.1) | 3210 (4.2) |
| Vaccinated with ≥1 dose | 2175 (73.7) | 46435 (95.3) | 1676 (88.4) | 39855 (95.6) | 3277 (90.9) | 72728 (95.8) |
| BNT162b2 (Pfizer-BioNTech) | 1552 (52.6) | 28576 (58.6) | 992 (52.3) | 18526 (44.4) | 1496 (41.5) | 30710 (40.4) |
| mRNA-1273 (Moderna) | 359 (12.2) | 7523 (15.4) | 227 (12.0) | 6870 (16.5) | 510 (14.1) | 11811 (15.6) |
| Mixed mRNA schedule | 264 (8.9) | 10336 (21.2) | 457 (24.1) | 14459 (34.7) | 1271 (35.2) | 30207 (39.8) |
| Bivalent booster<sup>c</sup> | NA | NA | NA | NA | 29 (0.8) | 679 (0.9) |
| **Interval in months between prior infection and specimen collection, median (IQR)** | 12 (11 – 14) | 13 (11 – 15) | 15 (4 – 17) | 4 (3 – 15) | 7 (4 – 17) | 7 (5 – 10) |
| **Interval in months between last vaccine dose and specimen collection, median (IQR)** | 4 (2 – 6) | 2 (1 – 4) | 4 (3 – 5) | 3 (2 – 5) | 5 (3 – 8) | 5 (3 – 7) |

Data are n (%) or median (Interquartile range)
Abbreviations: IQR, interquartile range; NA, not applicable; PI, prior infection
<sup>a</sup> Categories of comorbidity are not mutually exclusive
<sup>b</sup> BA.4/5 prior infections considered until September 6, 2022, 60 days before the end of the study period
<sup>c</sup> At least one bivalent booster dose (Pfizer-BioNTech or Moderna)

Across the analysis period, the proportion of cases and controls with a documented prior infection increased from 1.1% and 4.9%, respectively, during the BA.1 period, to 2.7% and 10.6% during the BA.2 period and 4.6% and 15.8% during the BA.4/5 period (**Table 2**). Overall, most prior infections were due to omicron including 53.2% among cases (134/252) and 61.9% among controls (11,648/18,822). The proportion of cases and controls with a prior pre-omicron infection was similar across BA.1 (1.1% and 4.8%), BA.2 (1.8% and 4.4%) and BA.4/5 analysis periods (1.4% and 4.0%). Conversely, the proportion of cases and controls with a prior omicron infection increased from the BA.1 (0.0% and 0.2%) to BA.2 (0.9% and 6.2%) and BA.4/5 (3.2% and 11.9%) periods. Somewhat counter-intuitively, the proportion with prior infections was lower among unvaccinated than among one- and two-dose vaccinated participants and was only slightly higher among unvaccinated than among three-dose vaccinated participants in all periods (**Supplementary figure 2**). During the BA.4/5 period, 626 (19.5%) of unvaccinated controls had a documented prior infection compared to 209 (35.1%) of those vaccinated with one dose, 1421 (23.4%) with two doses, and 9755 (14.8%) with ≥3 doses.

**Table 2.** Prior infection history and vaccination status in cases and controls stratified by omicron sublineage analysis period among elderly adults 60 years or older

| Prior infection and<br>Vaccination status | BA.1 period |  | BA.2 period |  | BA.4/5 period |  |
| --- | --- | --- | --- | --- | --- | --- |
|  | Cases<br>n=2951 | Controls<br>n=48724 | Cases<br>n=1897 | Controls<br>n=41702 | Cases<br>n=3607 | Controls<br>n=75938 |
| No prior infection | 2918 (98.9) | 46323 (95.1) | 1845 (97.3) | 37292 (89.4) | 3440 (95.4) | 63927 (84.2) |
| Unvaccinated | 772 (26.2) | 2154 (4.4) | 218 (11.5) | 1610 (3.9) | 316 (8.8) | 2584 (3.4) |
| 1 dose | 72 (2.4) | 477 (1.0) | 23 (1.2) | 263 (0.6) | 33 (0.9) | 386 (0.5) |
| 2 doses | 1099 (37.2) | 11110 (22.8) | 197 (10.4) | 3286 (7.9) | 329 (9.1) | 4648 (6.1) |
| 3 doses | 964 (32.7) | 31804 (65.3) | 1136 (59.9) | 21432 (51.4) | 1030 (28.6) | 19325 (25.4) |
| 4 doses | 11 (0.4) | 778 (1.6) | 271 (14.3) | 10701 (25.7) | 1460 (40.5) | 30533 (40.2) |
| 5 doses | NA | NA | NA | NA | 272 (7.5) | 6451 (8.5) |
| Prior pre-omicron infection | 32 (1.1) | 2322 (4.8) | 35 (1.8) | 1840 (4.4) | 51 (1.4) | 3012 (4.0) |
| Unvaccinated | 4 (0.1) | 133 (0.3) | 2 (0.1) | 104 (0.2) | 7 (0.2) | 154 (0.2) |
| 1 dose | 5 (0.2) | 177 (0.4) | 4 (0.2) | 86 (0.2) | 1 (0.0) | 122 (0.2) |
| 2 doses | 14 (0.5) | 990 (2.0) | 13 (0.7) | 480 (1.2) | 5 (0.1) | 440 (0.6) |
| 3 doses | 9 (0.3) | 969 (2.0) | 13 (0.7) | 840 (2.0) | 19 (0.5) | 1003 (1.3) |
| 4 doses | 0 (0.0) | 53 (0.1) | 3 (0.2) | 330 (0.8) | 16 (0.4) | 1043 (1.4) |
| 5 doses | NA | NA | NA | NA | 3 (0.1) | 250 (0.3) |
| Prior omicron infection | 1 (0.0) | 79 (0.2) | 17 (0.9) | 2570 (6.2) | 116 (3.2) | 8999 (11.9) |
| Unvaccinated | 0 (0.0) | 2 (0.0) | 1 (0.1) | 133 (0.3) | 7 (0.2) | 472 (0.6) |
| 1 dose | 0 (0.0) | 0 (0.0) | 1 (0.1) | 36 (0.1) | 3 (0.1) | 87 (0.1) |
| 2 doses | 1 (0.0) | 32 (0.1) | 1 (0.1) | 525 (1.3) | 12 (0.3) | 981 (1.3) |
| 3 doses | 0 (0.0) | 45 (0.1) | 10 (0.5) | 1229 (2.9) | 35 (1.0) | 2961 (3.9) |
| 4 doses | 0 (0.0) | 0 (0.0) | 4 (0.2) | 647 (1.6) | 48 (1.3) | 3541 (4.7) |
| 5 doses | NA | NA | NA | NA | 11 (0.3) | 957 (1.3) |

Data are n (%)
Abbreviations: NA, not applicable

The percentage of controls with repeat vaccine (booster) doses increased during the three periods (**Table 2**). By the time of specimen collection, 67.4% and <0.1% of controls had received up to three or four doses during the BA.1 period; 56.4% and 26.8% during the BA.2 period; and 30.7% and 44.1% during BA.4/5 period, when 679 (<1%) had received a bivalent booster dose.

During BA.1, BA.2 and BA.4/5 periods, two- vs. four-dose estimated VE against hospitalisation among those without documented prior infection was: 78% (95%CI:75-80) vs. 96% (95%CI:93-98); 60% (95%CI:50-97) vs. 84% (95%CI:81-87); and 40% (95%CI:30-49) vs. 68% (95%CI:63-72), respectively, consistent with longer median time since second vs fourth dose (e.g. 13 vs. 4 months for BA.4/5) (**Figure 2** and **Supplementary table 1**). VE estimates among those without prior infection decreased with more recent sublineages and was improved with more doses, notably against BA.4/5 hospitalisation, after standardizing for time since vaccination (**Table 3**). For example, at <3 months post-vaccination, two- vs. three-dose VE was 86% (95%CI:78-91) vs. 93% (95%CI:92-94) during BA.1, 66% (95%CI:28-84) vs. 85% (95%CI:81-88) during BA.2, and 55% (95%CI:-95, 89) vs. 82% (95%CI:68-90) during BA.4/5 periods. At 3-5 months post-vaccination, two- vs. three-dose VE was 78% (95%CI:74-81) vs. 87% (95%CI:84-89) during BA.1, 46% (95%CI: 12-66) vs. 73% (95%CI:68-77) during BA.2, and 40% (95%CI: -5-66) vs. 67% (95%CI:48-78) during BA.4/5 periods. Among infection-naïve adults, there were no differences in three vs. four dose VE at <3 months post-vaccination during BA.1 (93% vs. 96%), BA.2 (85% vs. 82%) or BA.4/5 periods (82% vs. 85%) nor at 3-5 months post-vaccination during the BA.4/5 period for three (67%), four (64%) or five (57%) dose recipients, recognizing wide confidence intervals for some of these estimates (**Table 3**).

**Figure 2.**
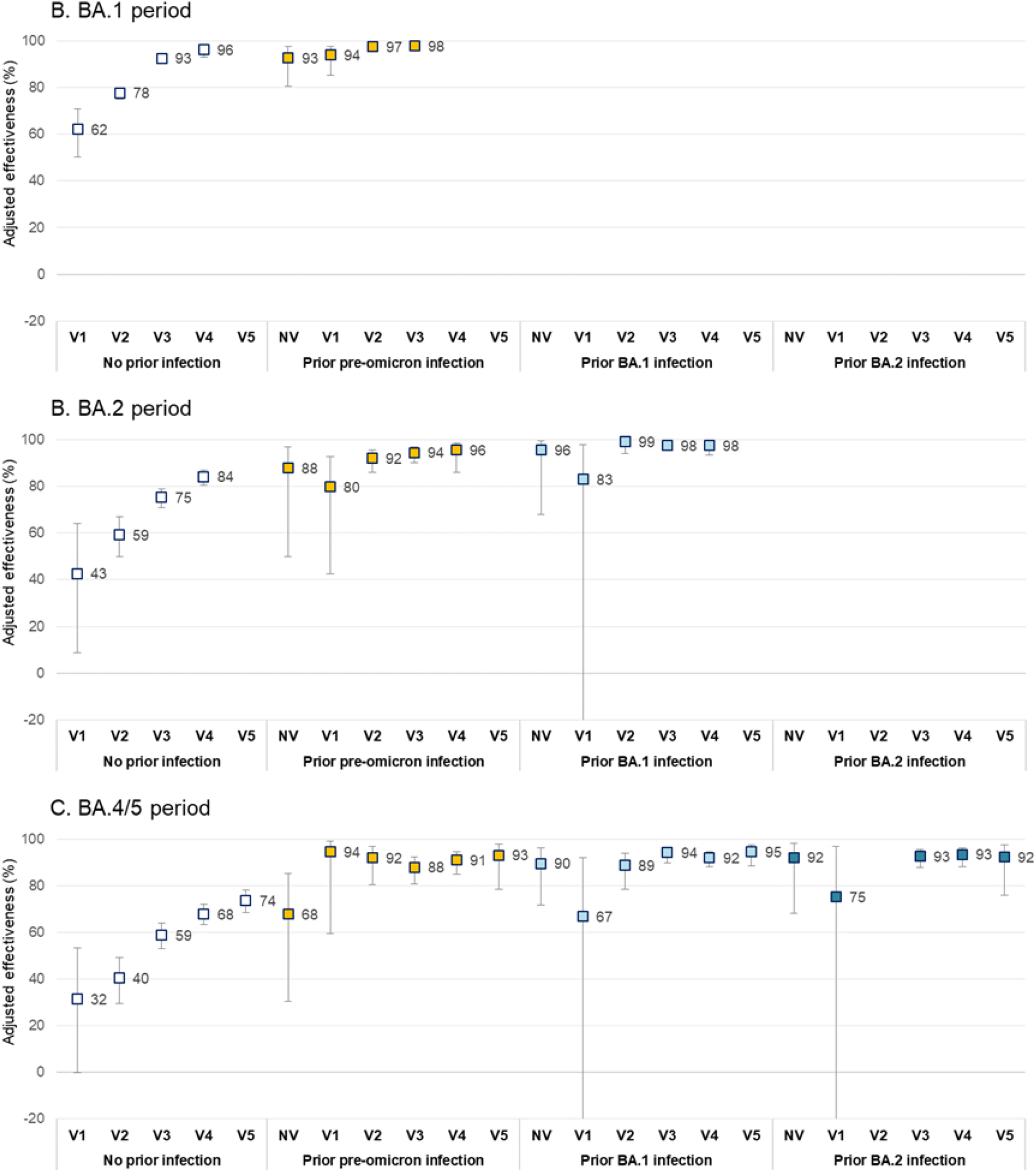
Estimated protection against omicron-related hospitalisations conferred by prior pre-omicron or omicron infection with or without vaccination by omicron sublineage analysis period (effectiveness and 95% confidence intervals) Note: Logistic regression models adjusted for sex, age (60-69, 70-79, 80-89 and ≥90 years old), origin (home, private homes for elderly, other), epidemiological week, multimorbidity (≥2 conditions), chronic respiratory disease, chronic heart disease, cancer, obesity, immunosuppressive condition, neurological disease.

**Table 3.** Vaccine and hybrid protection against omicron-related hospitalisations by time since last immunogenic event (vaccination or primary infection)

|  | **Adjusted effectiveness<sup>a</sup>, % (95% confidence intervals)** |  |  |  |  |
| --- | --- | --- | --- | --- | --- |
| **Interval since vaccination or time since primary infection** | **<3 months<sup>b</sup>** | **3 to 5 months** | **6 to 8 months** | **9 to 11 months** | **12 to 14 months** |
| **BA.1 period** |  |  |  |  |  |
| **1. When vaccination is last event:** |  |  |  |  |  |
| **Vaccination without PI** |  |  |  |  |  |
| 2 doses | 86 (78, 91) | 78 (74, 81) | 76 (73, 79) | 68 (37, 84) | NA |
| 3 doses | 93 (92, 94) | 87 (84, 89) | NA | NA | NA |
| 4 doses | 96 (92, 98) | NA | NA | NA | NA |
| **Vaccination after PI** |  |  |  |  |  |
| 2 to 4 doses and pre-omicron PI | 95 (92, 97) | 98 (94, 99) | 96 (90, 99) | NA | NA |
| **2. When PI is last event:** |  |  |  |  |  |
| **PI without vaccination** |  |  |  |  |  |
| Pre-omicron PI | NE | NE | NE | NE | NE |
| **PI after vaccination** |  |  |  |  |  |
| 2 to 4 doses and pre-omicron PI | NE | NE | NE | NE | NE |
| **BA.2 period** |  |  |  |  |  |
| **1. When vaccination is last event:** |  |  |  |  |  |
| **Vaccination without PI** |  |  |  |  |  |
| 2 doses | 66 (28, 84) | 46 (12, 66) | 58 (43, 69) | 61 (50, 70) | 73 (-13, 94) |
| 3 doses | 85 (81, 88) | 73 (68, 77) | 66 (48, 78) | NA | NA |
| 4 doses | 82 (78, 86) | 85 (69, 93) | NA | NA | NA |
| **Vaccination after PI** |  |  |  |  |  |
| 2 to 4 doses and pre-omicron PI | 95 (88, 98) | 92 (85, 95) | 84 (54, 94) | 87 (46, 96) | NA |
| 2 to 4 doses and omicron PI | 96 (90, 98) | NA | NA | NA | NA |
| **2. When PI is last event:** |  |  |  |  |  |
| **PI without vaccination** |  |  |  |  |  |
| Pre-omicron PI | NE | NE | NE | NE | NE |
| Omicron PI | NE | NE | NE | NE | NE |
| **PI after vaccination** |  |  |  |  |  |
| 2 to 4 doses and pre-omicron PI | NE | NE | NE | NE | NE |
| 2 to 4 doses and omicron PI | 95 (88, 98) | 99 (95, 100) | NA | NA | NA |
| **BA.4/5 period** |  |  |  |  |  |
| **1. When vaccination is last event:** |  |  |  |  |  |
| **Vaccination without PI** |  |  |  |  |  |
| 2 doses | 55 (-95, 89) | 40 (-5, 66) | 40 (5, 62) | 36 (17, 51) | 47 (35, 57) |
| 3 doses | 82 (68, 90) | 67 (60, 74) | 56 (49, 62) | 56 (45, 66) | NA |
| 4 doses | 80 (76, 83) | 64 (58, 69) | 52 (38, 63) | 75 (-7, 94) | NA |
| 5 doses | 73 (67, 78) | 57 (11, 79) | NA | NA | NA |
| **Vaccination after PI** |  |  |  |  |  |
| 2 to 5 doses and pre-omicron PI | 95 (90, 98) | 89 (81, 93) | 90 (79, 95) | 82 (54, 93) | NA |
| 2 to 5 doses and omicron PI | 94 (90, 96) | 94 (90, 97) | 95 (65, 99) | NA | NA |
| **2. When PI is last event:** |  |  |  |  |  |
| **PI without vaccination** |  |  |  |  |  |
| Pre-omicron PI | NE | NE | NE | NE | NE |
| Omicron PI | 90 (30, 99) | 93 (71, 98) | 84 (57, 94) | NA | NA |
| **PI after vaccination** |  |  |  |  |  |
| 2 to 5 doses and pre-omicron PI | NE | NE | NE | NE | NE |
| 2 to 5 doses and omicron PI | 91 (85, 95) | 94 (90, 96) | 92 (86, 96) | 88 (51, 97) | NA |

Note: Logistic regression models adjusted for sex, age (60-69, 70-79, 80-89 and $\geq 90$ years old), origin (home, private homes for elderly, other), epidemiological week, multimorbidity ( $\geq 2$ conditions), chronic respiratory disease, chronic heart disease, cancer, obesity, immunosuppressive condition, neurological disease.
<sup>b</sup> 7 to 90 days when vaccination is last event and 60 to 90 days when primary infection is last event
Abbreviations: NA, not applicable considering variant circulations or vaccine recommendations; PI, prior infection

When compared with infection-naïve individuals who had received their second dose ≥6 months earlier and were thus eligible for a booster dose, the relative VE of third and fourth doses was 70% (95%CI:67-73) and 85% (95%CI:72-92), respectively, during BA.1, 38% (95%CI:26-48) and 61% (95%CI:52-68) during BA.2, and 31% (95%CI:22-40) and 47% (95%CI:39-53) during BA.4/5 analysis periods (**Supplementary table 1**).

During BA.1, BA.2 and BA.4/5 periods, estimated effectiveness of pre-omicron vs. omicron infection alone (without vaccination) against hospitalisation was: 93% (95%CI:80-97) vs. [not estimable]; 88% (95%CI:50-97) vs. 96% (95%CI:68-99); and 69% (95%CI:30-85) vs. 90% (95%CI:79-95) (**Figure 2** and **Supplementary table 1**). Although not estimable for earlier periods or for prior pre-omicron infection, prior omicron infection alone was associated with a BA.4/5 hospitalisation risk reduction of 90% (95%CI: 30-99) at <3 months (days 60-89), 93% (95%CI: 90-97) at 3 to 5 months and 84% (95%CI: 57-94) at 6 to 8 months post-specimen collection (**Table 3**).

Hybrid immunity including prior infection and at least two vaccine doses was associated with higher protection against hospitalisation than vaccination alone during all analysis periods, and higher than pre-omicron infection alone during the BA.4/5 period (**Figure 2**). Regardless of the number of vaccine doses (2-5) or prior infection type (pre-omicron or omicron), hybrid immunity was associated with a hospitalisation risk reduction of ∼90% or more. Among individuals with hybrid immunity, booster doses were not associated with improved protection compared to two doses during any of the analysis periods (**Supplementary table 1**).

When vaccination was the last immunological event, hybrid protection associated with prior pre-omicron infection (among those who had received 2-5 vaccine doses) decreased slightly and non-significantly with time since last dose from 95% (95%CI:90-98) at <3 months, an interval after which fewer (1.1%) controls were twice vaccinated, to 82% (95%CI:54-93) at 9-11 months, a period after which more (51.4%) controls were twice vaccinated (**Table 3**). However, when prior infection was omicron, estimated hybrid protection during the 8 months after the last vaccine dose remained stable at ≥94% (longer follow-up not feasible). When a prior omicron infection was the last immunological event (not estimable for pre-omicron infection), hybrid protection (2-5 vaccine doses) also remained stable: 91% (95%CI:85-95) at <3 months (days 60-89), 94% (95%CI:90-96) at 3 to 5 months and 92% (95%CI:86-96) at 6 to 8 months.

In sensitivity analyses, results were similar when reinfection was defined with a 90-day interval (**Supplementary table 2**), with restriction to community-dwelling individuals (**Supplementary table 3**) and with exclusion of immunosuppressed individuals (**Supplementary table 4**).

## DISCUSSION

In this study, we show that among elderly adults, monovalent mRNA VE against omicron hospitalisation was lower for BA.4/5 than BA.1 or BA.2 sublineages, and was modified foremost by prior infection history rather than number of vaccine doses. Among infection-naïve elderly adults, VE estimates did not differ for three vs. four doses at <3 months post-vaccination for any sublineage, nor at 3-5 months post-vaccination during the BA.4/5 period for recipients of 3-5 doses. Infection-induced immunity in unvaccinated individuals was associated with better and longer lasting protection against hospitalisation than vaccination alone, slightly lower with prior infection due to pre-omicron than omicron. Whereas protection waned with time since either of these immunological events alone, hybrid immunity among those with both prior infection and at least two vaccine doses was more robust, exceeding 90% for at least 6-8 months regardless of type of prior infection or number of doses received.

Among infection-naïve elderly adults, estimated VE against omicron hospitalisation exceeded 90% for BA.1, was ∼85% for BA.2, and ∼80% for BA.4/5 during the 3 months after the last booster (third or fourth) dose, dropping by ∼15% after 3-5 months. These findings are consistent with observations elsewhere although previous studies did not assess all of these omicron sublineages and/or lacked simultaneous stratification for prior infection history. In preprint manuscripts among adults ≥18 years with unknown prior infection status, three-dose VE in Portugal was 93% against BA.2 and 77% against BA.4/5 hospitalisation^24^ and in the United States was 98% for BA.1, 82% for BA.2 and 72% for BA.4/5 hospitalisation.^8^ Our findings suggest that additional booster doses of monovalent mRNA vaccine did not gradually increase protection to new heights but rather reset protection to the levels achieved shortly after a previous dose. This conceptualization is also supported by two previous studies where VE against BA.4/5 hospitalisation was the same shortly after a third or fourth dose among adults without prior documented infection (66%) (preprint),^9^ and among adults with unknown prior infection (60%)^25^.

The substantial protection we observed among unvaccinated elderly adults and in association with prior infection alone corroborates and extends results from the meta-analysis by Bobrovitz et al. showing previous infection reduced the subsequent risk of severe COVID-19 by 83% at 3 months and 75% at 12 months during BA.1 and BA.2 dominant periods (preprint).^10^ In Qatar, pre-omicron and omicron infection without vaccination were associated, respectively, with a 36% and 69% reduced risk of symptomatic BA.4/BA.5 infection,^26^ but no study assessed protection against severe outcomes. We identified no other publication that assessed hybrid protection against BA.4/5-associated hospitalisation, but the Bobrovitz et al. meta-analysis reported that hybrid immunity including two or three vaccine doses reduced the risk of severe BA.1 and BA.2 by >95% at both the 3- or 6-month follow-up, similar to our findings for these sublineages (and also for BA.4/5).^10^

In our study, outcome ascertainment was comprehensive and specific for COVID-19-related hospitalisations. Cases and controls were elderly adults tested in a single type of facility for the same clinical indication, increasing comparability and reducing possible selection bias.^27^ Our study nevertheless has several limitations most likely on balance to result in under-estimation. As prior infection was documented through NAAT detection, some undiagnosed pre-omicron infections were missed, but many omicron infections were either undiagnosed or confirmed by self-performed (but undocumented) RADT, which became the predominant tests used from January 2022 even if NAAT access for older adults was maintained. Among our BA.4/5 controls, we identified 15% with a prior infection compared to 40% in the same age group in sero-prevalence studies conducted in July-August 2022 in Quebec and British Columbia, Canada.^13,28^ Missed prior infections in the reference group of unvaccinated “uninfected” individuals would result in underestimation of the protection induced by prior infection and hybrid immunity. VE among “uninfected” individuals would be underestimated if the proportion of missed prior infections among vaccinated individuals was smaller than the unvaccinated “uninfected” group, but would be overestimated if their proportion of missed prior infections was higher.

The unexpectedly low proportion (19%) of prior infections in unvaccinated controls compared to controls vaccinated with ≥2 doses during the BA.4/5 period may have been due to more missed infections or fewer exposure opportunities among unvaccinated older adults who may have more rigorously self-isolated. Both explanations would underestimate the protection from infection- and/or vaccine-induced immunity. Depending upon the magnitude of misclassification, the disproportionately lower rates of prior infection among unvaccinated adults may tend also to obscure the incremental value of additional doses, notably among those considered previously uninfected. Despite adjustment for age, sex, epidemiological week and comorbidities, residual confounding is possible. We did not have information on the use of Ritonavir-boosted Nirmatrelvir (Paxlovid) available in Quebec since mid-January 2022 for immunosuppressed adults and since mid-March 2022 for individuals ≥60 years with <2 vaccine doses and those up-to-date with vaccination but at higher risk of COVID-19 complications due to comorbidity.^29^ Since this medication may also reduce the risk of hospitalisation, failure to take it into account may have also tended to underestimate VE. Although our main analyses included immunocompromised individuals with potentially reduced response to vaccination, they represented only 7% of cases and controls and similar estimates were obtained with their exclusion. Our findings do not apply to bivalent vaccines or against less severe COVID-19. Extrapolation to newly circulating sublineages requires caution. Despite these limitations, our data align with and confirm observations from other immunological and epidemiological studies,^4-11^ while our analysis of three sublineages simultaneously, notably including BA.4/5, updates understanding and enables within lineage comparison as well as cross-reference with previous studies of vaccine and hybrid immunity against BA.1 and BA.2 only.

Our findings have methodological as well as clinical implications. Given the substantial effect modification by prior infection and vaccination, estimates of protection should be separately considered for each of these strata (vaccine only, prior infection only, both) rather than averaged as a global VE finding adjusted for prior infection as a confounder.^30^ This also applies to estimates of protection by sublineage and time since the last dose. For public health authorities and expert advisory committees, the effect modification by prior infection means that recommendations for additional vaccine doses may need to take into account the evolving proportion of the population already previously infected. Among previously-infected individuals, the plateauing of hybrid protection after two doses calls into question the incremental value of additional booster doses in preventing COVID-19 hospitalisation, whereas among the infection-naïve, waning vaccine protection over time may better justify booster doses.

## Conclusion

Elderly adults with history of both prior SARS-CoV-2 infection and at least two vaccine doses appear well-protected for a prolonged period against omicron hospitalisation, including BA.4/5. As such, the risk of COVID-19 hospitalisation foremost varies in older adults based on prior infection history and booster dose recommendations should take this important consideration into account. Immunisation programs aiming to prevent hospitalisations should prioritize efforts to administer booster doses to infection-naïve individuals who still constitute about half of the elderly adult population in high income countries by autumn 2022.

## Data Availability

The databases used in this study are a property of the Ministère de la Santé et des Services sociaux du Québec that was shared with the researchers under the legal mandate of the National Director of Public Health of Quebec under the Public Health Act, precluding data sharing with a third party. Aggregate data are available within the manuscript and the supplementary material.

## Acknowledgments

This work was supported by the Ministère de la santé et des services sociaux du Québec. We thank Louis Rochette (Institut National de Santé Publique du Québec) for his assistance in building the original databases used for the analyses, and Ève Dube and Denis Hamel (Institut National de Santé Publique du Québec) for sharing and discussing data of the populational surveys on the attitudes and behaviors of Quebec adults in relation to COVDI-19.

## Data sharing statement

The databases used in this study are a property of the “Ministère de la santé et des services sociaux du Québec” that was shared with the researchers under the legal mandate of the National Director of Public Health of Quebec under the Public Health Act, precluding data sharing with a third party. Aggregate data are available within the manuscript and the supplementary material.

