## Supplemental material for "Prior infection- and/or vaccine-induced protection against Omicron BA.1, BA.2 and BA.4/BA.5-related hospitalisations in older adults: a test-negative case-control study in Quebec, Canada"

**Supplementary table 1. Estimated protection against Omicron hospitalisations conferred by pre-Omicron or Omicron prior infection with or without vaccination, by Omicron sublineage analysis period**

| Period and exposure | Days since last event (vaccination or PI), median (IQR) | Estimated protection, % (95% confidence intervals) | | | |
|---|---|---|---|---|---|
| | | Compared to unvaccinated individuals without prior infection | | Compared to individuals vaccinated with 2 doses ≥6 months earlier | |
| | | Unadjusted | Adjusted | Unadjusted | Adjusted |
| **BA.1 period** | | | | | |
| No prior infection | | | | | |
| Unvaccinated | NA | Reference | Reference | NA | NA |
| 1 dose | 159 (50-270) | 58 (45, 68) | 62 (50, 71) | NA | NA |
| 2 doses | 192 (173-212) | 72 (69, 75) | 78 (75, 80) | Reference | Reference |
| 3 doses | 51 (30-74) | 92 (91, 92) | 93 (92, 93) | 71 (68, 74) | 70 (67, 73) |
| 4 doses | 21 (13-31) | 96 (93, 98) | 96 (93, 98) | 86 (75, 93) | 85 (72, 92) |
| Pre-Omicron PI | | | | | |
| Unvaccinated | 387 (288-564) | 92 (77, 97) | 93 (80, 97) | NA | NA |
| 1 dose | 238 (145-310) | 92 (81, 97) | 94 (85, 98) | NA | NA |
| 2 doses | 120 (76-196) | 96 (93, 98) | 97 (96, 99) | Reference | Reference |
| 3 doses | 63 (35-99) | 97 (95, 99) | 98 (96, 99) | 32 (-103, 78) | 4 (-219, 71) |
| **BA.2 period** | | | | | |
| No prior infection | | | | | |
| Unvaccinated | NA | Reference | Reference | NA | NA |
| 1 dose | 228 (111-362) | 35 (-1, 59) | 43 (9, 64) | NA | NA |
| 2 doses | 285 (244-314) | 56 (46, 64) | 60 (50, 67) | Reference | Reference |
| 3 doses | 117 (97-142) | 61 (54, 66) | 75 (71, 79) | 7 (-10, 21) | 38 (26, 48) |
| 4 doses | 31 (18-46) | 81 (78, 85) | 84 (81, 87) | 56 (46, 64) | 61 (52, 68) |
| Pre-Omicron PI | | | | | |
| Unvaccinated | 473 (229-525 | 86 (42, 97) | 88 (50, 97) | NA | NA |
| 1 dose | 326 (234-400) | 66 (6, 88) | 80 (42, 93) | NA | NA |
| 2 doses | 170 (138-252) | 80 (65, 89) | 92 (86, 96) | Reference | Reference |
| 3 doses | 105 (72-136) | 89 (80, 94) | 94 (90, 97) | 38 (-66, 77) | 39 (-75, 79) |
| 4 doses | 34 (20-50) | 93 (79, 98) | 96 (86, 99) | 63 (-48, 91) | 60 (-80, 91) |
| Omicron PI | | | | | |
| Unvaccinated | 101 (89-124) | 94 (60, 99) | 96 (68, 99) | NA | NA |
| 1 dose | 97 (63-120) | 80 (-50, 97) | 83 (-29, 98) | NA | NA |
| 2 doses | 100 (78-120) | 99 (90, 100) | 99 (94, 100) | Reference | Reference |
| 3 doses | 79 (50-103) | 94 (89, 97) | 98 (95, 99) | NE | NE |
| 4 doses | 28 (17-40) | 95 (88, 98) | 98 (93, 99) | NE | NE |
| **BA.4/5 period** | | | | | |
| No prior infection | | | | | |
| Unvaccinated | NA | Reference | Reference | NA | NA |
| 1 dose | 320 (228-468) | 30 (-2, 52) | 32 (0, 53) | NA | NA |
| 2 doses | 399 (353-440) | 42 (32, 51) | 40 (30, 49) | Reference | Reference |
| 3 doses | 227 (195-263) | 56 (50, 62) | 59 (53, 64) | 24 (14, 33) | 31 (22, 40) |
| 4 doses | 108 (78-142) | 61 (56, 66) | 68 (63, 72) | 32 (23, 40) | 47 (39, 53) |
| 5 doses | 34 (20-51) | 66 (59, 71) | 74 (68, 78) | 40 (29, 49) | 56 (48, 63) |
| Pre-Omicron PI | | | | | |
| Unvaccinated | 559 (345-600) | 63 (20, 83,) | 69 (30, 85) | NA | NA |
| 1 dose | 458 (346-532) | 93 (52, 99) | 94 (60, 99) | NA | NA |
| 2 doses | 302 (241-391) | 91 (77, 96) | 92 (80, 97) | Reference | Reference |
| 3 doses | 180 (115-235) | 85 (75, 90) | 88 (81, 92) | -87 (-453, 37) | -92 (-484, 37) |
| 4 doses | 94 (52-135) | 88 (79, 92) | 91 (85, 95) | -52 (-356, 50) | -34 (-319, 57) |
| 5 doses | 38 (24-53) | 90 (69 ,97) | 93 (78, 98) | -19 (-434, 74) | 13 (-328, 82) |
| Omicron PI | | | | | |
| Unvaccinated | 117 (115-224) | 88 (74, 94) | 90 (79, 95) | NA | NA |
| 1 dose | 157 (99-203) | 72 (10, 91) | 73 (12, 92) | NA | NA |
| 2 doses | 175 (110-222) | 90 (82, 94) | 91 (85, 95) | Reference | Reference |
| 3 doses | 121 (87-168) | 90 (86, 93) | 93 (90, 95) | 9 (-76, 53) | 13 (-71, 56) |
| 4 doses | 79 (49-108) | 89 (85, 92) | 92 (90, 94) | -5 (-98, 45) | 0 (-96, 49) |
| 5 doses | 36 (21-52) | 91 (83, 95) | 94 (88, 97) | 11 (-102, 61) | 18 (-103, 67) |

Note: Logistic regression models adjusted for sex, age (60-69, 70-79, 80-89 and ≥90 years old), origin (home, private homes for elderly, other), epidemiological week, multimorbidity (≥2 conditions), chronic respiratory disease, chronic heart disease, cancer, obesity, immunosuppressive condition, neurological disease.

Abbreviations: IQR, interquartile range; NA, not applicable; NE, not estimable; PI, prior infection

**Supplementary table 2. Estimated protection against Omicron hospitalisations conferred by prior infection with or without vaccination by Omicron sublineage analysis using a 90-day interval to define reinfections**

| | Estimated protection, % (95% confidence intervals) | | | | | |
|---|---|---|---|---|---|---|
| | BA.1 period (n=51465) | | BA.2 period (n=43029) | | BA.4/5 period (n=78547) | |
| **Exposure** | **Unadjusted** | **Adjusted** | **Unadjusted** | **Adjusted** | **Unadjusted** | **Adjusted** |
| No prior infection | | | | | | |
| Unvaccinated | Reference | Reference | Reference | Reference | Reference | Reference |
| 1 dose | 58 (45, 68) | 62 (51, 71) | 35 (-1, 59) | 43 (9, 64) | 30 (-2, 52) | 32 (0, 53) |
| 2 doses | 72 (69, 75) | 78 (75, 80) | 56 (46, 64) | 59 (50, 67) | 42 (32, 51) | 40 (30, 49) |
| 3 doses | 92 (91, 92) | 93 (92, 93) | 61 (54, 66) | 75 (71, 79) | 56 (50, 62) | 59 (53, 64) |
| 4 doses | 96 (93, 98) | 96 (93, 98) | 81 (78, 85) | 84 (81, 87) | 61 (56, 66) | 68 (63, 72) |
| 5 doses | NA | NA | NA | NA | 66 (59, 71) | 74 (68, 78) |
| Pre-Omicron PI | | | | | | |
| Unvaccinated | 91 (75, 97) | 92 (79, 97) | 86 (42, 97) | 88 (50, 97) | 63 (20, 83) | 68 (31, 85) |
| 1 dose | 92 (81, 97) | 94 (85, 98) | 66 (6, 88) | 80 (43, 93) | 93 (52, 99) | 94 (60, 99) |
| 2 doses | 96 (93, 98) | 97 (95, 98) | 80 (65, 89) | 92 (86, 96) | 91 (77, 96) | 92 (80, 97) |
| 3 doses | 98 (95, 99) | 98 (96, 99) | 89 (80, 94) | 94 (90, 97) | 85 (75, 90) | 88 (81, 92) |
| 4 doses | NA | NA | 93 (79, 98) | 96 (86, 99) | 88 (79, 92) | 91 (85, 95) |
| 5 doses | NA | NA | NA | NA | 90 (69, 97) | 93 (78, 98) |
| Omicron PI | | | | | | |
| Unvaccinated | NA | NA | NE | NE | 88 (72, 95) | 90 (77, 96) |
| 1 dose | NA | NA | 74 (-95, 96) | 78 (-72, 97) | 79 (13, 95) | 79 (13, 95) |
| 2 doses | NA | NA | NE | NE | 90 (82, 95) | 91 (84, 95) |
| 3 doses | NA | NA | 96 (90, 98) | 98 (95, 99) | 91 (87, 94) | 93 (90, 95) |
| 4 doses | NA | NA | 95 (86, 98) | 97 (92, 99) | 90 (85, 93) | 93 (90, 95) |
| 5 doses | NA | NA | NA | NA | 91 (83, 95) | 94 (88, 97) |

Note: Logistic regression models adjusted for sex, age (60-69, 70-79, 80-89 and ≥90 years old), origin (home, private homes for elderly, other), epidemiological week, multimorbidity (≥2 conditions), chronic respiratory disease, chronic heart disease, cancer, obesity, immunosuppressive condition, neurological disease.

Abbreviations: NA, not applicable considering variant circulations or vaccine recommendations; NE, not estimable; PI, prior infection

**Supplementary table 3. Estimated protection against Omicron hospitalisations conferred by pre-Omicron or Omicron prior infection with or without vaccination by Omicron sublineage analysis period among community-dwelling elderly adults 60 years or older aged 60 years or older**

| | **Estimated protection, % (95% confidence intervals)** | | | | | |
|---|---|---|---|---|---|---|
| | **BA.1 period (n=38133)** | | **BA.2 period (n=33434)** | | **BA.4/5 period (n=59972)** | |
| **Exposure** | **Unadjusted** | **Adjusted** | **Unadjusted** | **Adjusted** | **Unadjusted** | **Adjusted** |
| No prior infection | | | | | | |
| Unvaccinated | Reference | Reference | Reference | Reference | Reference | Reference |
| 1 dose | 61 (47, 71) | 61 (48, 72) | 64 (32, 81) | 65 (34, 81) | 27 (-9, 50) | 29 (-6, 52) |
| 2 doses | 74 (72, 77) | 80 (77, 82) | 61 (51, 69) | 60 (50, 68) | 42 (31, 51) | 39 (27, 49) |
| 3 doses | 94 (93, 95) | 94 (93, 95) | 74 (69, 78) | 77 (72, 81) | 55 (48, 60) | 56 (49, 62) |
| 4 doses | 96 (92, 98) | 96 (93, 98) | 85 (81, 88) | 83 (79, 87) | 62 (57, 67) | 68 (63, 72) |
| 5 doses | NA | NA | NA | NA | 66 (59, 73) | 72 (66, 78) |
| Pre-Omicron PI | | | | | | |
| Unvaccinated | 92 (75, 98) | 93 (78, 98) | 82 (25, 96) | 84 (34, 96) | 73 (26, 90) | 77 (37, 92) |
| 1 dose | 96 (83, 99) | 97 (86, 99) | 78 (10, 95) | 83 (27, 96) | 91 (32, 99) | 92 (38, 99) |
| 2 doses | 98 (94, 99) | 98 (95, 99) | 91 (72, 97) | 91 (73, 97) | 90 (72, 96) | 90 (74, 96) |
| 3 doses | 98 (95, 99) | 98 (95, 99) | 95 (86, 99) | 96 (87, 99) | 85 (72, 92) | 86 (73, 93) |
| 4 doses | NA | NA | 96 (69, 99) | 96 (71, 99) | 85 (70, 92) | 87 (75, 93) |
| 5 doses | NA | NA | NA | NA | 81 (23, 95) | 86 (41, 97) |
| Omicron PI | | | | | | |
| Unvaccinated | NA | NA | 93 (48, 99) | 94 (56, 99) | 87 (71, 94) | 90 (76, 95) |
| 1 dose | NA | NA | 71 (-114, 96) | 66 (-158, 96) | 69 (-28, 92) | 66 (-40, 92) |
| 2 doses | NA | NA | 98 (84, 100) | 98 (86, 100) | 88 (78, 94) | 89 (80, 94) |
| 3 doses | NA | NA | 96 (86, 99) | 96 (87, 99) | 88 (82, 92) | 91 (86, 94) |
| 4 doses | NA | NA | 96 (71, 99) | 96 (72, 100) | 90 (83, 94) | 92 (86, 95) |
| 5 doses | NA | NA | NA | NA | 90 (59, 98) | 91 (63, 98) |

Note: Logistic regression models adjusted for sex, age (60-69, 70-79, 80-89 and ≥90 years old), epidemiological week, multimorbidity (≥2 conditions), chronic respiratory disease, chronic heart disease, cancer, obesity, immunosuppressive condition, neurological disease.

Abbreviations: NA, not applicable considering variant circulations or vaccine recommendations; PI, prior infection

**Supplementary table 4. Estimated protection against Omicron hospitalisations conferred by prior infection with or without vaccination by Omicron sublineage analysis period restricted to elderly adults 60 years or older without immunosuppression**

| | Estimated protection, % (95% confidence intervals) | | | | | |
|---|---|---|---|---|---|---|
| | BA.1 period (n=44073) | | BA.2 period (n=38210) | | BA.4/5 period (n=70096) | |
| **Exposure** | **Unadjusted** | **Adjusted** | **Unadjusted** | **Adjusted** | **Unadjusted** | **Adjusted** |
| No prior infection | | | | | | |
| Unvaccinated | Reference | Reference | Reference | Reference | Reference | Reference |
| 1 dose | 57 (44, 67) | 61 (49, 70) | 33 (-6, 58) | 41 (4, 63) | 30 (-4, 52) | 28 (-7, 51) |
| 2 doses | 74 (71, 77) | 79 (76, 81) | 55 (45, 63) | 59 (49, 67) | 43 (32, 51) | 40 (29, 50) |
| 3 doses | 93 (92, 94) | 93 (92, 94) | 62 (55, 68) | 76 (71, 80) | 57 (51, 63) | 59 (53, 64) |
| 4 doses | NE | NE | 82 (78, 85) | 84 (80, 87) | 62 (57, 67) | 69 (64, 73) |
| 5 doses | NA | NA | NA | NA | 70 (64, 75) | 78 (73, 82) |
| Pre-Omicron PI | | | | | | |
| Unvaccinated | 91 (76, 97) | 93 (80, 97) | 85 (38, 96) | 87 (48, 97) | 60 (13, 81) | 66 (26, 84) |
| 1 dose | 95 (84, 98) | 96 (88, 99) | 64 (0, 87) | 78 (37, 92) | 92 (44, 99) | 93 (52, 99) |
| 2 doses | 96 (93, 98) | 97 (95, 98) | 78 (61, 88) | 92 (85, 95) | 92 (78, 97) | 93 (81, 97) |
| 3 doses | 98 (94, 99) | 98 (95, 99) | 88 (78, 93) | 94 (89, 97) | 86 (77, 92) | 89 (82, 94) |
| 4 doses | NA | NA | 92 (69, 98) | 95 (80, 99) | 87 (78, 93) | 91 (84, 95) |
| 5 doses | NA | NA | NA | NA | 91 (61, 98) | 94 (74, 98) |
| Omicron PI | | | | | | |
| Unvaccinated | NA | NA | 94 (58, 99) | 95 (66, 99) | 89 (75, 95) | 91 (79, 96) |
| 1 dose | NA | NA | 79 (-54, 97) | 84 (-26, 98) | 80 (20, 95) | 80 (19, 95) |
| 2 doses | NA | NA | 98 (89, 100) | 99 (94, 100) | 91 (83, 95) | 92 (85, 96) |
| 3 doses | NA | NA | 95 (89, 98) | 98 (96, 99) | 91 (87, 94) | 94 (90, 96) |
| 4 doses | NA | NA | 93 (82, 98) | 97 (91, 99) | 88 (84, 92) | 92 (89, 94) |
| 5 doses | NA | NA | NA | NA | 87 (76, 93) | 92 (84, 96) |

Note: Logistic regression models adjusted for sex, age (60-69, 70-79, 80-89 and ≥90 years old), origin (home, private homes for elderly, other), epidemiological week, multimorbidity (≥2 conditions), chronic respiratory disease, chronic heart disease, cancer, obesity, neurological disease.

Abbreviations: NA, not applicable considering variant circulations or vaccine recommendations; NE, not estimable; PI, prior infection

**Supplementary figure 1. Study Population**

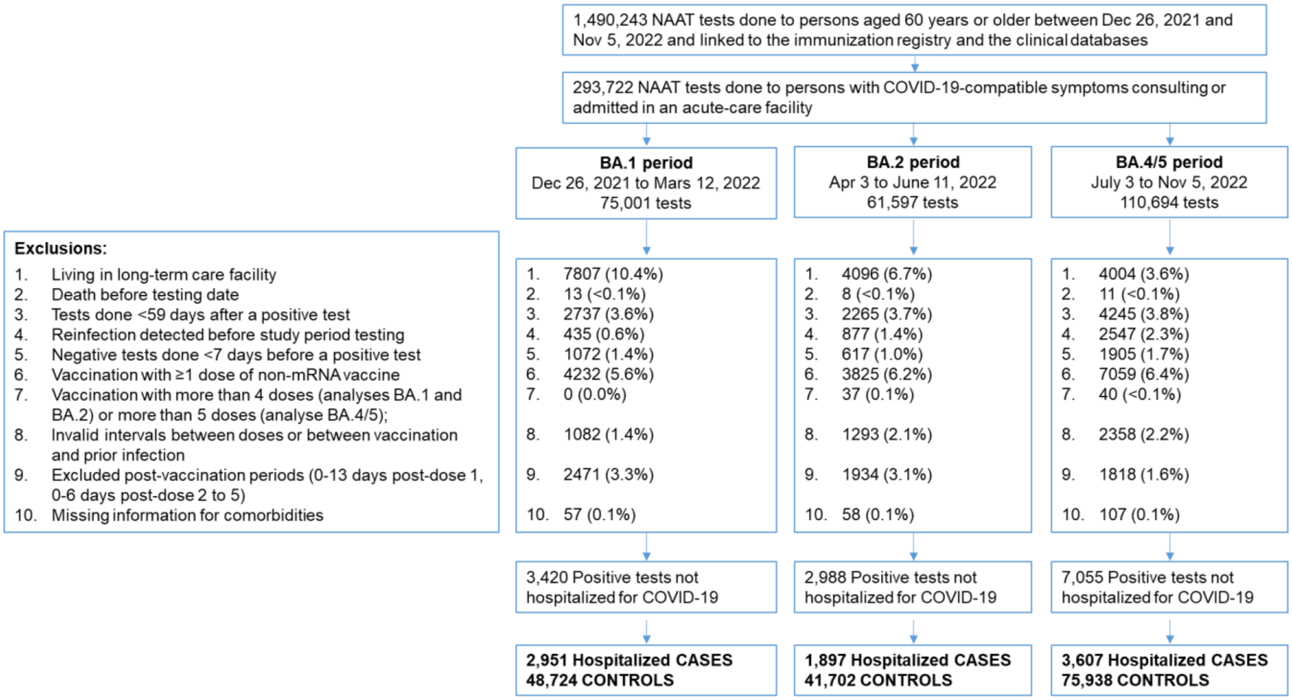

Note: Subvariant periods do not include periods with mixed circulation

Abbreviations: NAAT, nucleic acid amplification testing

**Supplementary figure 2. Proportion of controls with detected prior infection by vaccination status and Omicron sublineage analysis period**

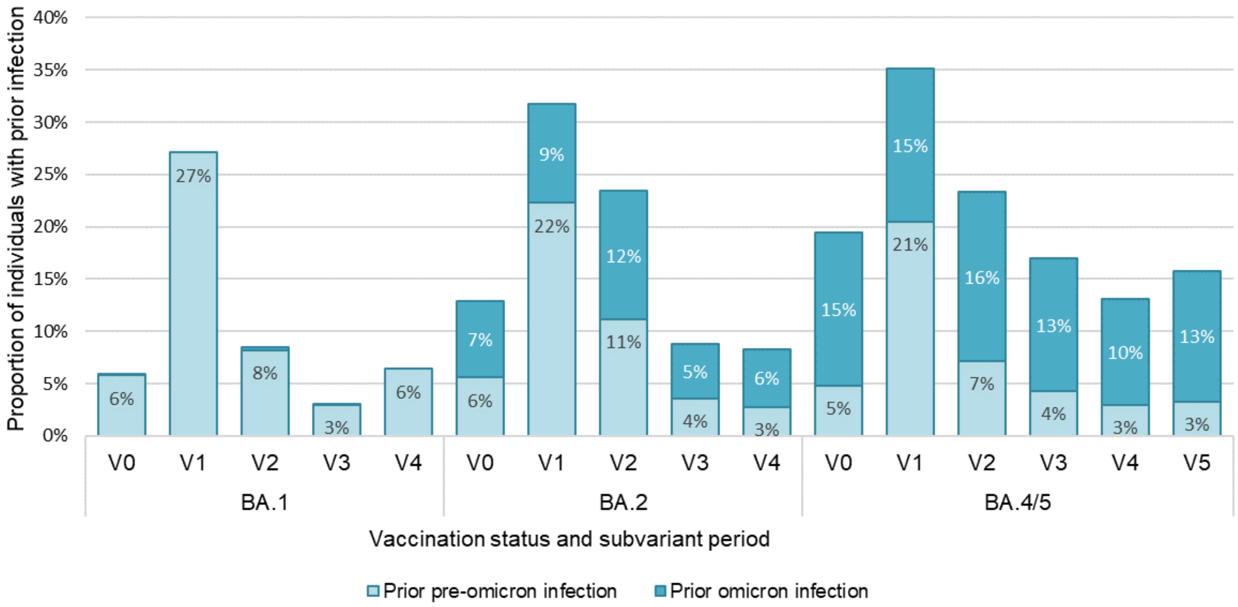

Abbreviations: V0, unvaccinated; V1, one dose; V2, two doses; V3, three doses; V4, four doses; V5, five doses
